# Losing ground at the wrong time: Trends in self-reported influenza vaccination uptake in Switzerland, Health Survey 2007–2017

**DOI:** 10.1101/2020.06.05.20123026

**Authors:** Kathrin Zürcher, Marcel Zwahlen, Claudia Berlin, Matthias Egger, Lukas Fenner

**Author notes:** **Correspondence:** Dr. Lukas Fenner, Institute of Social and Preventive Medicine (ISPM), University of Bern, Mittelstrasse 43, CH-3012 Bern, Switzerland.

## Abstract

**Objectives:** We studied time trends in seasonal influenza vaccination and associations with socioeconomic and health-related determinants in Switzerland, overall and in people aged ≥ 65 years.

**Design:** Three cross-sectional surveys.

**Participants:** Individuals who participated in the Swiss Health Surveys 2007, 2012, and 2017. We calculated the proportion reporting influenza vaccination in the last 12 months, and performed multivariable logistic regression analyses.

**Results:** The proportion of reporting a history of influenza vaccination overall was 31.9% (95% confidence intervals [95% CI] 31.4–32.4); and dropped from 34.5% in 2007 to 28.8% in 2017. The uptake of vaccination within the past 12 months was 16% in 2007 and similar in 2012 and 2017 (around 14%). In people with chronic disease, uptake dropped from 43.8% in 2007 to 37.1% in 2012 and to 31.6% in 2017 (p<0.001). In people aged ≥ 65 years, uptake dropped from 47.8% in 2007 to 38.5% in 2012 to 36.2% in 2017 (p<0.001). Similarly, a decrease in vaccine uptake was seen in people with poor self-reported health status (39.4%, 33.1%, and 27.0%). In logistic regression, self-reported vaccination coverage decreased in the 65 to 75 years old (adjusted odds ratio (aOR) aOR 0.56, 95% Cl 0.48–0.66 between 2007 and 2012; aOR 0.89, 95% CI 0.77–1.03). Uptake was positively associated with the ≥ 65 age group, living in French-speaking and urban areas, history of smoking, bad self-reported health status, private/semiprivate health insurance, having a medical profession, and having any underlying chronic disease. Use of any alternative medicine therapy was negatively associated with influenza vaccination (aOR 0.72, 95% CI 0.67–0.80).

**Conclusion:** Influenza vaccination coverage was low in older and chronically ill persons. Significant efforts are required in preparing for the flu season 2020/21 to reduce the double burden of COVID-19 and seasonal influenza. These efforts should include campaigns but also novel approaches using social media.

**Strengths and limitations of this study:**

- Data analysis of the Swiss Health Survey 2007, 2012, and 2017 focussing on influenza vaccine uptake overall and in the age group ≥65 years in Switzerland.
- The Swiss Health Survey is a nationwide, representative survey that is repeated every five years using the same methodology.
- Analyses were weighted and adjusted for a wide range of important cofactors.
- We calculated percent of people reporting having been vaccinated and associations between vaccination status and socio-demographic and health-related factors.
- Influenza vaccination status is self-reported in the Swiss Health Survey and the reliability of the data not ascertained.

## Introduction

Seasonal influenza is pandemic, and a challenge for surveillance, control and treatment (1). Worldwide, it causes 3 to 5 million cases of severe illness each year and kills 250,000 to 500,000 people (2), particularly infants, the elderly, and the chronically ill. In Switzerland, influenza is responsible for 111,000 to 331,000 medical consultations yearly and 1,000 to 5,000 hospitalizations (3). The current COVID-19 pandemic shows the impact of respiratory viruses on the burden of infectious diseases and the importance of vaccines in the control of viral respiratory diseases (4, 5).

In 2003, the World Health Assembly adopted a resolution urging member states to reach a target for uptake of influenza vaccines of 75% among people at high risk by 2010 (6). The Federal Office of Public Health (FOPH) in Switzerland has vaccine recommendations in place since 2007, which target mainly elderly people, but also those with chronic illnesses (including children older than six months), premature infants, pregnant women, residents of long-term health care facilities and those are in regular contact with vulnerable populations (7).

We earlier analyzed the data from the Swiss Health Survey 2007 and 2012 and showed that overall influenza vaccine uptake in Switzerland decreased from 2007 to 2012 (8). To examine recent trends and associations of socio-demographic characteristics and health-related factors with influenza vaccination practices in Switzerland, we analyzed the data from the most recent nationally representative health survey, in 2017, and compared the results with those from 2012 and 2007.

## Material and methods

### Survey sample

The cross-sectional Swiss Health Survey has been conducted every five years since 1992 by the Swiss Federal Statistical Office (SFSO). The survey is a multistage probability sample drawn from all residents not living in institutions in Switzerland (8, 9). Conducted between January and December of the year, the survey collects data using computer-assisted telephone interviews and self-completed questionnaires.

We compared the data set from 2017 with the data from 2012 and 2007 and only used survey data from the written forms. We excluded the telephone interviews because they were shorter and did not include all the questions as the written forms. We included a total of 51,582 people who responded to a written questionnaire; 14,393 responded in 2007, 18,357 in 2012, and 18,632 in 2017.

### Data collection and definitions

All three surveys included two identical questions about influenza vaccination: (i) Have you ever had an influenza vaccination? (answers: yes, no, unknown). (ii) If yes: When were you last vaccinated? (answer: date).

The questionnaire collected demographic and socioeconomic as well as health-related information (Table 1) (5) on chronic diseases such as diabetes, cancer, lung, cerebrovascular, and cardiovascular disease. The basic health insurance is mandatory in Switzerland and cover for illness, maternity and accidents and offers the same range of services to all insured people (10). The respondents’ health insurance plan regarding coverage in case of hospitalization (private, semi-private or general ward), free choice of physicians and coverage of complementary medicine (including acupuncture, traditional Chinese medicine, homoeopathy, and osteopathy) in the past 12 months was recorded as well.

**Table 1.**
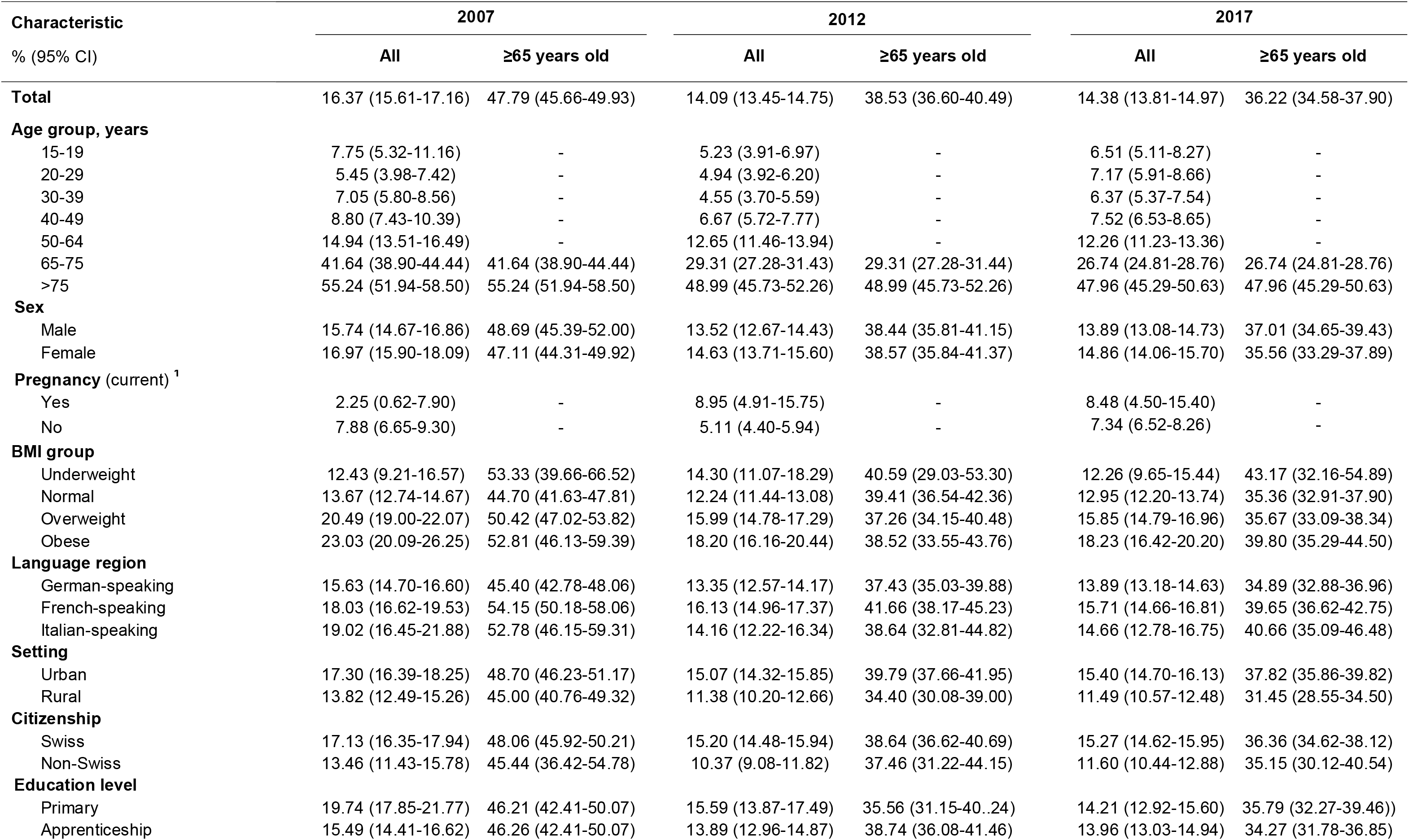
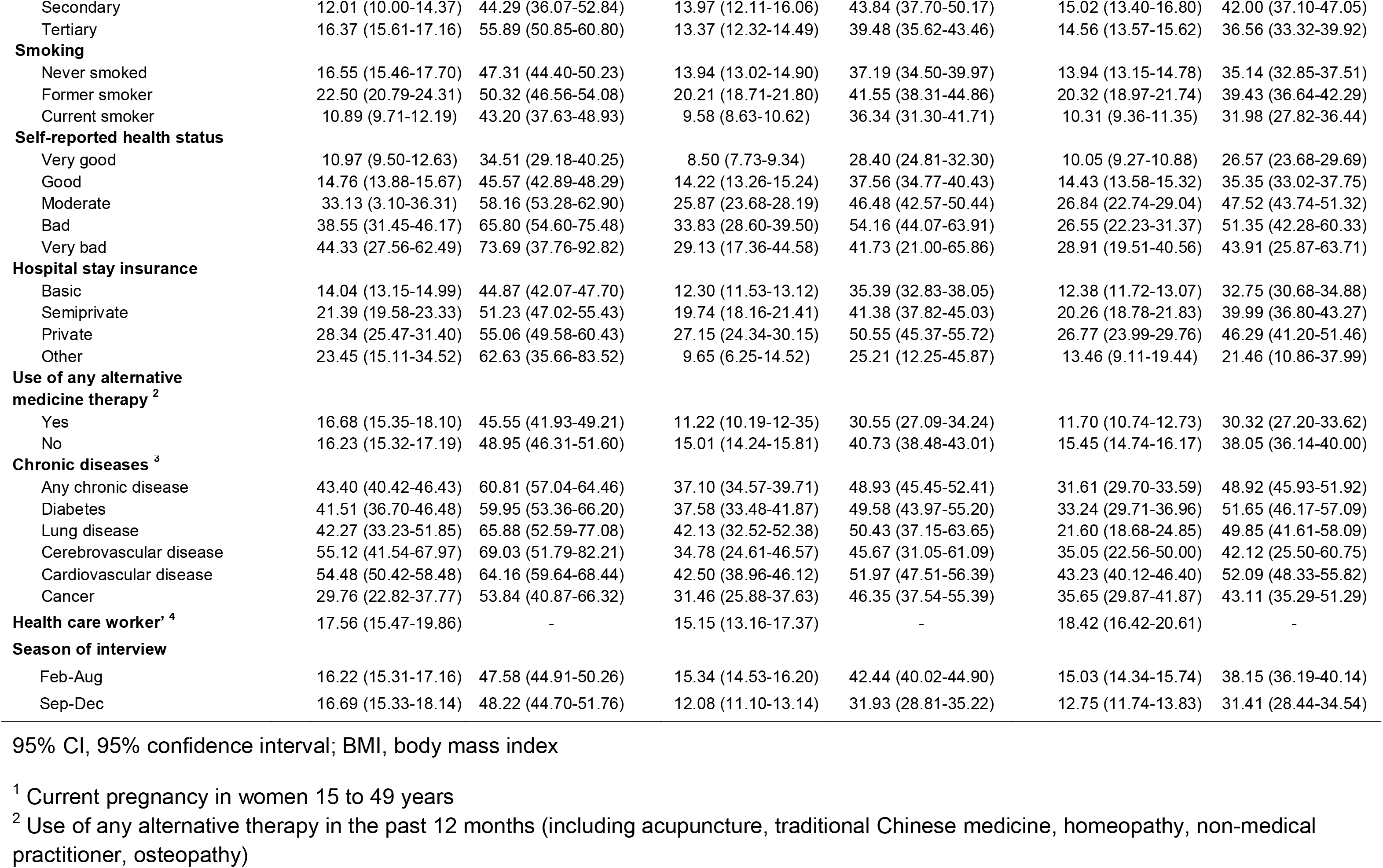
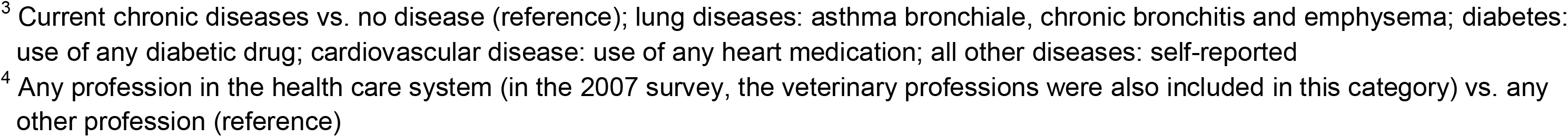
Percent of people reporting having been vaccinated for influenza in the last 12 months in Switzerland, 2012 and 2017, overall and among people ≥65 years old.

Pregnancy was recorded as current pregnancy among women 15 to 49 years old. Current chronic conditions included asthma, chronic bronchitis and emphysema. Diabetes was defined by the use of any diabetic drug, cardiovascular disease by the use of any heart medication, and all other chronic diseases were recorded as self-reported. We defined any chronic disease as the presence of at least one of the mentioned diseases. Health care workers were defined as individuals reporting profession in the health care system.

Cantons are the administrative subdivisions of Switzerland (see Figure 1).

**Figure 1.**
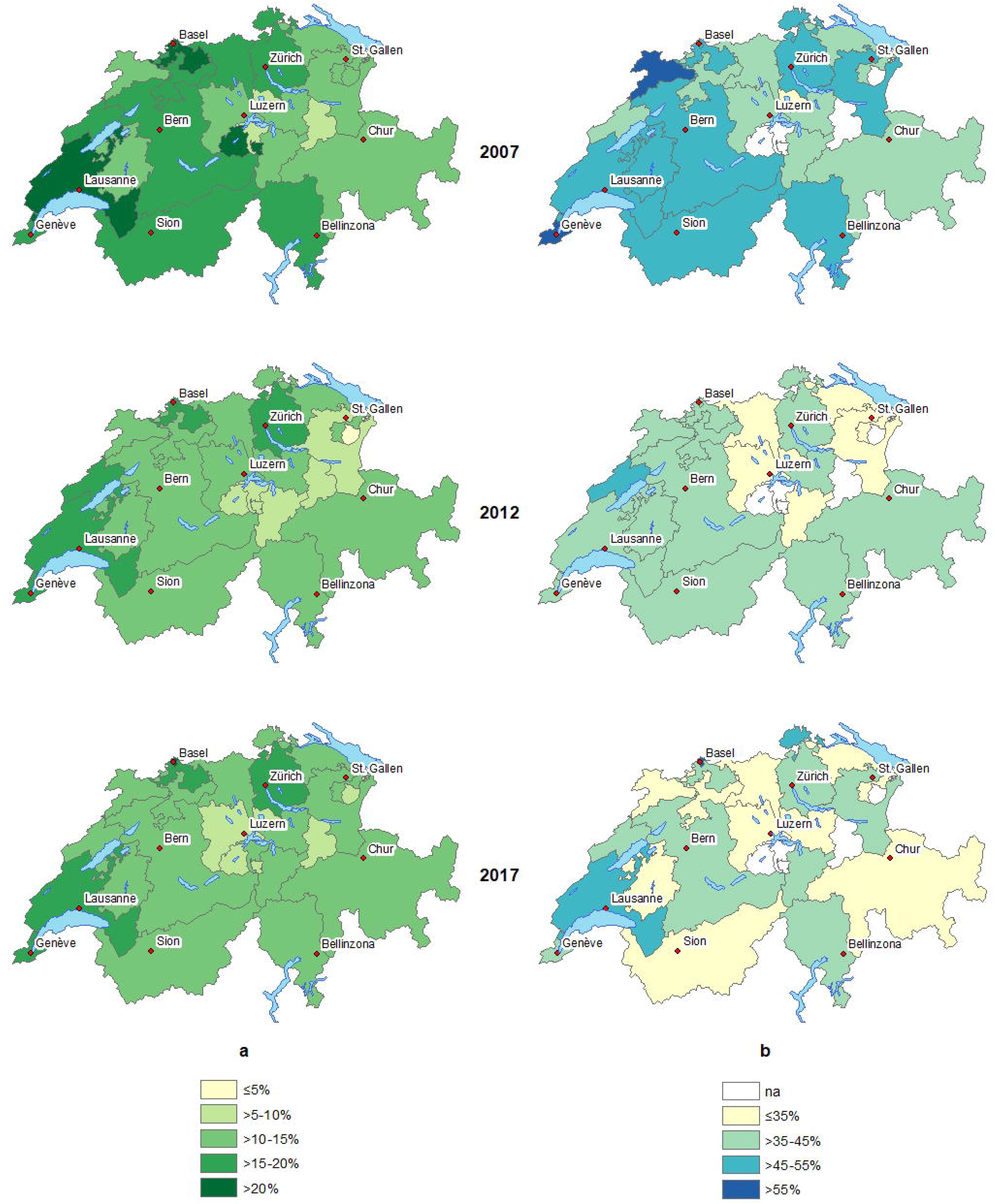
Geographical distribution (by canton) of people reporting influenza vaccination in the last 12 months in Switzerland, 2012 and 2017. Panel a, overall; panel b, people aged ≥ years. na, not applicable (no. of respondents <100).

### Statistical analysis

For each of the three survey years we calculated the proportions (overall and ≥65 years) that reported having been vaccinated within the last 12 months. We estimated associations between vaccination status and socio-demographic and health-related factors. We included an interaction term with the year of survey and the variable of interest in multivariable logistic regression models. We used the SFSO’s survey weights and reported all proportions and unadjusted and adjusted odds ratio (ORs and aOR) with the corresponding 95% confidence intervals (95% CI) derived from robust standard error calculations (Table 2). All analyses were performed in Stata (version 15.1, Corporation, College Station, Texas, USA).

**Table 2.** Associations of having been vaccinated for influenza in the last 12 months with socio-demographic characteristics and health-related factors (as compared to no vaccination) in Switzerland, 2012 and 2017, overall and among people ≥65 years old.

| Characteristic | All |  | ≥65 years old |  |
| --- | --- | --- | --- | --- |
|  | Adjusted OR (95% CI) | P value | Adjusted OR (95% CI) | P value |
| **Year of survey** |  | <0.001 |  | <0.001 |
| 2007 | 1 |  | 1 |  |
| 2012 | 0.74 (0.66-0.83) |  | 0.60 (0.51-0.71) |  |
| 2017 | 0.72 (0.64-0.80) |  | 0.57 (0.49-0.66) |  |
| **Age group, years** |  | <0.001 |  | <0.001 |
| 15-19 | 1.07 (0.67-1.71) |  | - |  |
| 20-29 | 1 |  | - |  |
| 30-39 | 1.05 (0.77-1.44) |  | - |  |
| 40-49 | 1.32 (0.99-1.75) |  | - |  |
| 50-64 | 2.27 (1.74-2.96) |  | - |  |
| 65-75 | 6.26 (4.78-8.20) |  | 1 |  |
| >75 | 12.94 (9.81-17.08) |  | 2.04 (1.82-2.30) | <0.001 |
| **Sex** |  | 0.66 |  | 0.37 |
| Male | 1 |  | 1 |  |
| Female | 0.97 (0.88-1.08) |  | 1.06 (0.93-1.21) |  |
| **Pregnancy** |  | 0.67 |  |  |
| Yes | 0.91 (0.56-1.70) |  | - |  |
| No | 1 |  | - |  |
| **BMI group** |  | 0.87 |  | 0.93 |
| Underweight | 1 |  | 1 |  |
| Normal | 0.97 (0.74-1.28) |  | 0.88 (0.58-1.34) |  |
| Overweight | 0.94 (0.72-1.25) |  | 0.89 (0.59-1.36) |  |
| Obese | 0.99 (0.74-1.32) |  | 0.91 (0.58-1.41) |  |
| **Language region** |  |  |  |  |
| German-speaking | 1 | <0.001 | 1 | 0.010 |
| French-speaking | 1.31 (1.20-1.44) |  | 1.22 (1.07-1.38) |  |
| Italian-speaking | 0.95 (0.82-1.12) |  | 1.05 (0.86-1.29) |  |
| **Setting** |  | <0.001 |  | 0.017 |
| Urban | 1.24 (1.12-1.38) |  | 1.19 (1.03-1.37) |  |
| Rural | 1 |  | 1 |  |
| **Citizenship** |  | 0.80 |  | 0.199 |
| Swiss | 1 |  | 1 |  |
| Non-Swiss | 0.98 (0.85-1.13) |  | 0.87 (0.70-1.08) |  |
| **Education level** |  | <0.001 |  | 0.064 |
| Primary | 1 |  | 1 |  |
| Apprenticeship | 1.00 (0.88-1.14) |  | 1.05 (0.89-1.23) |  |
| Secondary | 1.11 (0.93-1.31) |  | 1.14 (0.91-1.43) |  |
| Tertiary | 1.26 (1.09-1.46) |  | 1.26 (1.04-1.53) |  |
| **Smoking** |  | <0.001 |  | 0.009 |
| Never smoked | 1 |  | 1 |  |
| Former smoker | 1.17 (1.05-1.28) |  | 1.22 (1.07-1.39) |  |
| Current smoker | 0.91 (0.81-1.02) |  | 1.01 (0.85-1.20) |  |
| **Self-reported health status** |  | <0.001 |  | <0.001 |
| Very good | 1 |  | 1 |  |
| Good | 1.16 (1.03-1.31) |  | 1.30 (1.10-1.53) |  |
| Moderate | 1.56 (1.36-1.80) |  | 1.65 (1.36-2.01) |  |
| Bad | 2.13 (1.70-2.65) |  | 2.03 (1.46-2.81) |  |
| Very bad | 2.07 (1.26-3.40) |  | 1.79 (0.92-3.45) |  |
| **Hospital stay insurance** |  | <0.001 |  | <0.001 |
| Basic | 1 |  | 1 |  |
| Semiprivate | 1.43 (1.30-1.59) |  | 1.42 (1.24-1.63) |  |
| Private | 1.78 (1.56-2.03) |  | 1.86 (1.56-2.21) |  |
| Other | 1.28 (0.86-1.92) |  | 0.89 (0.50-1.58) |  |
| **Use of any alternative medicine therapy**<sup>1</sup> |  | <0.001 |  | <0.001 |
| Yes | 0.72 (0.66-0.80) |  | 0.65 (0.57-0.75) |  |
| No |  |  | 1 |  |
| **Chronic diseases**<sup>2</sup> |  |  |  |  |
| Any chronic disease | 1.84 (1.69-2.09) | <0.001 | 1.68 (1.51-1.88) | <0.001 |
| Diabetes | 1.54 (1.33-1.77) | <0.001 | 1.46 (1.23-1.73) | <0.001 |
| Lung disease | 1.81 (1.46-2.26) | <0.001 | 1.59 (1.16-2.18) | 0.004 |
| Cerebrovascular disease | 1.12 (0.74-1.69) | 0.59 | 0.91 (0.58-1.42) | 0.679 |
| Cardiovascular disease | 1.50 (1.34-1.68) | <0.001 | 1.44 (1.26-1.64) | <0.001 |
| Cancer | 1.22 (0.97-1.55) | 0.09 |  |  |
| **Health care worker**<sup>3</sup> | 1.92 (1.64-2.24) | <0.001 | - |  |

95% CI, 95% confidence interval; BMI, body mass index; OR, odds ratio
Model adjusted for all variables included in the table. Unadjusted ORs are shown in Supplementary Table S2.
<sup>1</sup> Current pregnancy in women 15 to 49 years
<sup>2</sup> Use of any alternative therapy in the past 12 months (including acupuncture, traditional Chinese medicine, homeopathy, non-medical practitioner, osteopathy)
<sup>3</sup> Any current chronic disease versus no chronic disease (reference); lung diseases: asthma bronchiale, chronic bronchitis and emphysema vs. no lung disease (reference); diabetes: use of any diabetic drug vs. no diabetes (reference); cardiovascular disease: use of any heart medication vs. no cardiovascular disease (reference); all other diseases: self-reported disease vs. no disease (reference)
<sup>4</sup> Estimates from separate models which included the variable “any chronic disease” instead of specific chronic diseases
<sup>5</sup> Any profession in the health care system (in the 2007 survey, the veterinary professions were also included in this category) vs. any other profession (reference)

We visualized changes in the frequency of vaccination uptake, and geographical distributions of the population that reported vaccination for influenza at the cantonal level using ArcGIS version 10.5 (Redlands, CA, USA).

### Ethics statement

Data were collected anonymized and ethical approval was not required but we obtained permission to analyze and publish the data through a contract with the SFSO (Ref. 624.110–1).

## Results

### Trends of influenza vaccinations status over time

The proportion of survey participants reporting a history of influenza vaccination overall was 31.9% (95% CI 31.4–32.4), having dropped from 34.5% in 2007 to 28.8% in 2017. The proportion reporting vaccination within the past 12 months was 16.4% (95% CI 15.6–17.2) in 2007, dropped to 14.1% (95% CI 13.5–14.8) in 2012, and remained at this level in 2017 (14.4%, 95% Cl 13.8–15.0, p<0.001, Table 1). Among those ≥ 65 years old, the principal target population of the Swiss recommendations (7), vaccination in the past 12 months dropped from 47.8% (95% Cl 45.7–49–9) in 2007, to 38.5% (95% CI 36.6–40.5) in 2012, and 36.2% (95% CI 34.6–37.9) in 2017 (p<0.001). For those with any chronic disease, another at-risk population, the frequency of influenza vaccination dropped from 43.8% (95% Cl 40.9–46.8) in 2007, to 37.1% (95% Cl 34.6–39.7) in 2012, and further down to 31.6% (95% Cl 29.7–33.6) in 2017. Similarly, a decrease in vaccine uptake was seen in people with poor self-reported health status (39.4%, 33.1%, and 27.0%, Figure 2).

**Figure 2.**
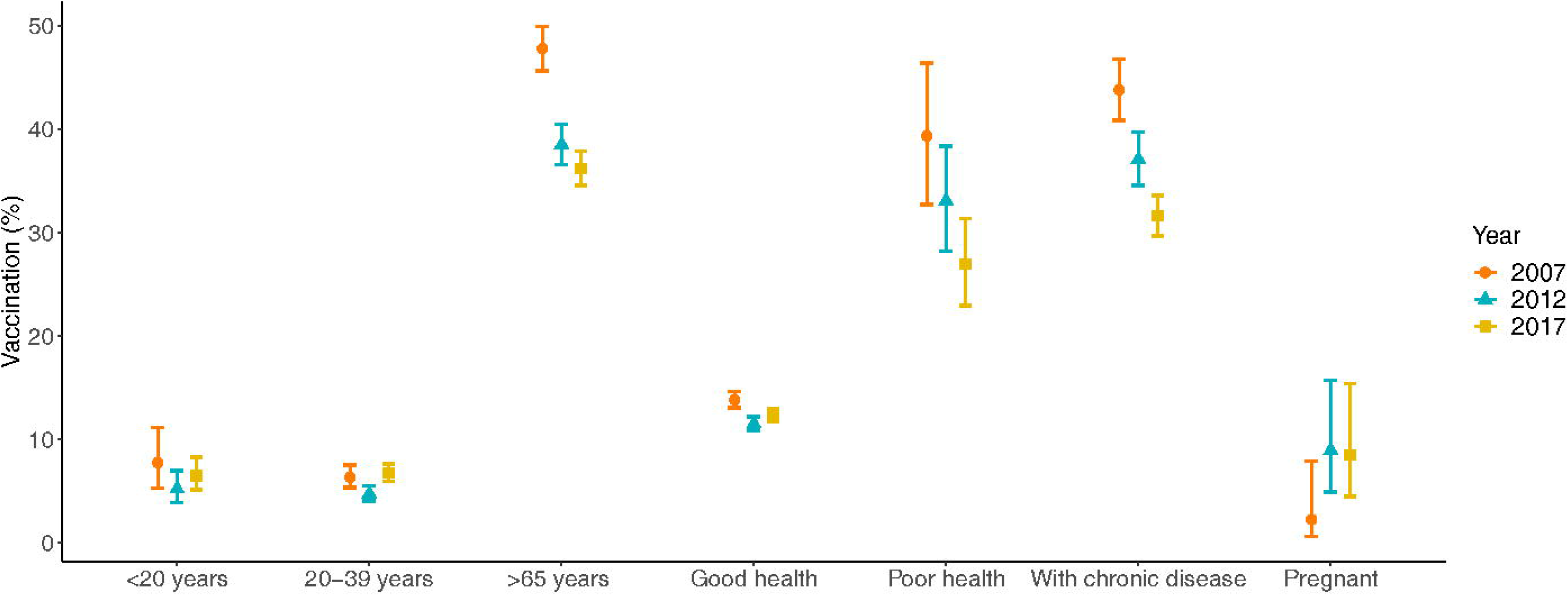
Temporal trends of selected groups of people reporting influenza vaccination in the last 12 months in Switzerland, comparing 2012 and 2017. Percent of people reporting having been vaccinated and the 95% confidence intervals are presented. Good health: self-reported health status good and very good; Poor health: self-reported health status bad and very bad; With chronic disease: any chronic illness

From 2007 to 2012 the self-reported influenza vaccination decreased in age group 15–19 years (aOR 0.51, 95% Cl 0.27–0.99) but increased from 2012 to 2017 in the younger age groups (e.g., aOR 1.59, 95% CI 0.92–2.74 in the 15 to 19 years age group; aOR 1.19. 96% Cl 0.84–1.69 in the 30 to 39 years age group, Figure 2). In contrast, it decreased in the 65 to 75 years old between 2007 and 2017 (aOR 0.56, 95% Cl 0.48–0.66 for 2007- 2012; aOR 0.89, 95% CI 0.77–1.03 for 2012–2017, Supplementary Table S1). The p-value from the test for an interaction between period and age group was 0.01 for 2007–2012 and 0.051 for 2012–2017. In pregnant women, an increase of influenza vaccination was observed between 2007 and 2012 (aOR 4.43, 95% CI 0.96–20.42, p = 0.02), with no further increase between 2012 and 2017 (aOR 0.94, 95% CI 0.36–2.47, p = 0.45). The temporal trends over the period 2007 to 2012 and 2012 to 2017 were not associated with age, language region, urban/rural setting, citizenship, use of complementary medicine, or type of hospital stay insurance (p>0.05, Supplementary Table S1),

### Influenza vaccination status in different population groups

In all three surveys, self-reported influenza vaccination was highest in the age group ≥65 years, and also higher in urban settings, among Swiss citizens, former smokers, and in persons with supplemental health insurance (Table 1). In all three surveys, the proportion of self-reported influenza vaccination was higher in the French- and Italian-speaking region compared to the German region. Persons with chronic diseases were more frequently vaccinated for influenza, with uptake ranging from around 20% to 57% depending on age group and type of chronic disease. A lower proportion of self-reported vaccination was observed in ages 15 years to 50 years (range 4–9%), the German-speaking area of Switzerland, current smokers, and person who reported their health status as very good. Pregnant women had increased frequency of influenza vaccination in 2007 and 2012, and then the influenza vaccine uptake stabilized between 2012 and 2017 (Table 1, Figure 2). The regional differences in the vaccination status in the last 12 months in the overall population and the ≥65 age group are shown in Figures 1A and 1B.

### Individual and health system factors associated with the influenza vaccination status

In all three surveys, influenza vaccination in the last 12 months was positively associated with age 65 years or older, living in French-speaking and urban areas, having a tertiary education, history of smoking, bad self-reported health status, private/semiprivate hospital stay insurance, being a health care worker, and having any underlying chronic disease. Use of any alternative medicine therapy was negatively associated with influenza vaccination (aOR 0.72, 95% CI 0.67–0.80). We found no association with sex, body mass index, or non-Swiss citizenship.(Table 2). Similar findings were observed when analyzing only participants’ ≥65 years old. Supplementary Table 2 shows unadjusted and adjusted ORs of the associations between self-reported vaccination for influenza in the last 12 months and sociodemographic characteristics and health-related factors.

## Discussion

Progress toward the WHO target of 75% vaccination coverage among high-risk groups has not only stalled in Switzerland, some of the gains made in earlier years were lost by 2017. Of particular concern is that the self-reported seasonal influenza vaccination rate among elderly persons declined from 47.8% in 2007 to 38.5% in 2012, and declined further to 36.2% in 2017. People at risk due to underlying chronic diseases reported coverage of influenza vaccination of 31.6% in 2017. Overall, after declining from 16.4% in 2007 to 14.1% in 2012, coverage in Switzerland was, with 14.4%, similar in 2017.

A decline in influenza vaccine coverage in recent years has been observed in other European countries. In the European Region of WHO, vaccine uptake in older people ranged from 0.03% to 76.3% over seven seasons (2008/2009–2014/2015). The median was 34.4%. In 2014/15, only Scotland reached the WHO and European Council goal with an uptake of 75%. Among countries providing data for seasons 2008/2009 and 2014/2015, over half reported a drop in vaccination coverage among older people (11). Among people with chronic diseases, coverage in the European Region ranged from 0.3% in Kyrgyzstan to 86.8% in Georgia in 2014/2015 (11).

The decline in coverage and an increase in vaccine hesitancy is of concern. In the last years, several countries of the WHO European Region including Switzerland, had large outbreaks of vaccine-preventable diseases, including measles, rubella and influenza. Switzerland and other countries identified steps to improve vaccination coverage for influenza and other infectious diseases (12, 13). The relationship between vaccination uptake, knowledge, attitudes, and awareness is complex (14). Reasons for people not getting vaccinated against influenza include underestimation of disease severity, fear of side effects, and the cost of vaccines (15–17). Often people are unaware that they should receive the vaccination (15). Of note, the coverage of measles vaccination increased to almost 90% in young adults in Switzerland, probably due to the campaigns and a national measles strategy (18). The experience with measles could serve as a model for influenza.

The advent of the novel coronavirus SARS-Cov-2 has profoundly changed everyday life in Switzerland and elsewhere. It is unclear how the COVID-19 pandemic will affect attitudes toward vaccines. The director of the US Centers for Disease Control has warned that a possible second wave of Covid-19 could be worse than the first (19). This has already been seen in the “Spanish flu” pandemic of 1918/19. The Spanish flu affected Switzerland in two waves. The first one occurred in July 1918, the second, more severe one, in October–November 1918 (20). However, even in the absence of a second wave, a resurgence of Covid-19 that coincides with the start of the flu season could significantly stress health care systems. An effective and safe vaccine against COVID 19 is unlikely to become widely available in 2020. Therefore, low influenza vaccination rates at around half the 75% coverage recommended for high-risk groups constitute a hazard that merits prompt, focused attention by public health authorities.

A concerted effort to increase influenza vaccination coverage in 2020/21, when the COVID-19 pandemic to continue to pose a threat to the public’s health, is urgently needed. Influenza vaccination prevents deaths, morbidity, hospital admissions, and other adverse health outcomes, in target populations such as older people, chronically ill people (21–26), and also children (27) and pregnant women (28). The continued promotion of infection control measures such as avoiding close contact with sick people, covering one’s nose and mouth while coughing or sneezing, social distancing and hand hygiene will contribute to reducing the spread of both influenza and Covid-19 (3).

Our study has several limitations. Influenza vaccination status is self-reported in the Swiss Health Survey, and the reliability of the data unclear. For example, vaccination coverage could be lower if social desirability bias led to an overestimation of uptake. Conversely, incomplete recall of vaccinations could bias coverage downwards. Individuals younger than 15 years are excluded from the survey, but coverage in this age group is probably even lower than in the 15 to 19-year-olds. A strength of our analysis is the fact that the survey is a nationwide and representative, and repeated every five years using the same methodology. Also, the analyses were weighted and adjusted for a range of potential confounders, which did not substantially change the results.

In conclusion, we need to increase influenza vaccination uptake, particularly in the elderly and chronically ill, who are also the risk groups most heavily affected by COVID-19. These efforts should include classic information campaigns, but novel approaches using social media should also be considered (29, 30). Recommendations by health care professionals are essential to improve influenza vaccination coverage, such as client reminder/recall and standing orders (31). The preparation of influenza season 2020/21 must start now to address the double burden of COVID-19 and seasonal influenza.

## Patient and Public involvement

The Swiss Health Survey is a nationwide, representative survey that is repeated every five years using the same methodology. The survey is conducted by the Swiss Federal Statistical Office (SFSO) on behalf of the Swiss Government. The content of the survey is based on a holistic and dynamic health framework and contains questions on essential topics for the public and politics. The wording of the questions is regularly harmonized with the statistical offices of other countries in Europe.

## Data Availability

Data may be obtained upon request from the Swiss Federal Statistical Office (Switzerland).

## Acknowledgements

We thank the Swiss Federal Statistical Office for providing the data of the Swiss Health Survey 2012 and 2017, and the people who participated in the surveys.

